# Temporal dynamics of the gut microbiome preceding celiac disease in genetically at-risk children

**DOI:** 10.1101/2025.05.29.25328357

**Authors:** Angelica P. Ahrens, Kristian Lynch, Heikki Hyöty, Richard E. Lloyd, Joseph Petrosino, Eric W. Triplett, Daniel Agardh

## Abstract

Longitudinal study of the microbial dysbiosis preceding celiac disease (CD) is needed, particularly in the first several years of life. Within the Environmental Determinants of Diabetes in the Young (TEDDY) multi-national prospective cohort study, a case-cohort study of 306 CD cases (i.e., seroconverting by 48 months of age), with controls matched 2:1 by site, gender, and time of birth, was assessed. Temporal microbiome case-control dynamics were modelled by 16S rRNA analysis of monthly sequential stool samples taken from age three months up to age four (or until the development of CD). Significant differences were identified across time, including key taxa that break down gluten and influence inflammation, all *before* the development of autoantibodies. Key bacterial associations with environmental factors such as diet were assessed using detailed longitudinal nutrient intake and diary data, along with genetic variants conferring high CD risk.

## INTRODUCTION

Celiac disease (CD) is a chronic immune-mediated disorder triggered by gluten ingestion in genetically predisposed individuals carrying the HLA-DQ2 and/or HLA-DQ8 haplotypes^1^. While genetic susceptibility is required for disease development, environmental factors play a key role in modulating immune responses to gluten and influencing disease onset^2^. Among these, dietary composition and gut microbiota have emerged as critical determinants of CD risk^3^.

Epidemiological studies suggest that the quantity of gluten introduction may influence CD risk. Data from The Environmental Determinants of Diabetes in the Young (TEDDY) Study^4^ indicates that higher gluten intake during the first years of life is associated with an increased risk of CD autoimmunity and progression to clinical disease ^5^. However, the mechanisms underlying this association remain unclear since exposure to other environmental factors also contribute to the disease risk^6^. Emerging evidence suggests that dietary fiber intake may reduce CD risk, possibly by shaping gut microbiota composition and promoting immune tolerance^7^. Fiber fermentation by gut microbes leads to the production of short-chain fatty acids (SCFAs), such as butyrate, which exert immunomodulatory effects and maintain epithelial integrity^8^. As with other autoimmune diseases such as T1D, the microbiome has been postulated to have a role in CD pathogenesis ^9^. At the phylum level, decreases in the abundance of *Firmicutes* species and increases in *Proteobacteria* species have been detected in both children and adults with active CD^10,11^. The *Proteobacteria* phylum includes several immuno-stimulatory taxa and has been associated with flares in other inflammatory diseases, such as Inflammatory Bowel Disease (IBD). Further studies have demonstrated that decreases in anti-inflammatory bacteria such as *Bifidobacterium* and increases in the proportion of *Bacteroides* and *Escherichia coli* have also been associated with active CD^12,13^. Other work suggests that the normal gut microbiome may be protective in reducing inflammation associated with gluten ingestion, and that certain microbial constituents may be involved in the development of CD^14^.

Other studies have reported distinct microbial signatures in individuals with CD, including a reduction in anti-inflammatory taxa (*Bifidobacterium* and *Faecalibacterium prausnitzii*) and an expansion of pro-inflammatory species (*Proteobacteria*)^15,16^. Importantly, microbial dysbiosis has been observed in genetically at-risk infants before the onset of CD^17^. Some gut bacteria, including *Lactobacillus* and *Bifidobacterium*, can metabolize gluten peptides and potentially reduce their immunogenicity, whereas others, such as *Pseudomonas aeruginosa*, may enhance gluten toxicity by modifying peptide digestion^18,19^. These findings support a complex interplay between diet, microbial metabolism, and immune activation in CD pathogenesis.

Although there is increasing evidence of strong linkage between human gut microbial communities and autoimmune disorders. Several reports indicate linkage of both viruses and microbiota with CD development^1–9^. In children at genetic risk, gastrointestinal infections increase the risk of CDA. HLA genotype, infant gluten consumption, breastfeeding, and rotavirus vaccination modify this risk, suggesting that a complex series of interactions between infectious agents, diet, and genetics promote this disease^10^. Even among those who carry the highest genetic susceptibility for CD^11^ (e.g., HLA-DR3-DQ2 homozygous individuals) and are exposed to gluten, only a minority will develop CD, suggesting that other factors must play critical roles. However, prior studies linking gut microbiome and virome with autoimmune diseases are limited in scale and scope and fail to provide consistent or persuasive linkages to specific etiologic agents or conclusions that certain bacteria or viruses are linked. In addition, very few prospective studies have evaluated the role of bacteria and viruses already before CDA and those that have are small in sample size focused on CD and those that have are small in sample size. In this case-cohort study, nested in the prospective TEDDY birth cohort, we investigate the relationship between early-life dietary patterns, gut microbiome composition, and the risk of developing CD. By analyzing a longitudinal, genetically at-risk cohort, we aim to identify microbial and dietary factors that could play a role prior providing novel insights into CD etiology.

## RESULTS

This TEDDY ancillary study represents the largest multinational prospective investigation of the CD microbiome to date, with quarterly stool sampling from 3 to 48 months and longitudinal CDA assessments, confirmed by biopsy-proven CD diagnosis. For this study’s case-control cohort, a total of 137 CD cases and 232 randomly chosen controls were included. For each case, sample collection ceased once CDA was detected (**Fig. 1a**), while controls were sampled continuously until 48 months. The median age at seroconversion was 28.3 months (IQR: 21.8-36.7). CD cases were further stratified by age at seroconversion: early (≤24 months, median 18.5), mid (24 to ≤36 months; median 29.2), and late (36 to ≤48 months; median 42.0) seroconversion, respectively (**Table 1**). The number of stool samples from CD cases and number of CD cases available for this study as a function of the age in months at time of CDA seroconversion (**Fig. 1b**).

**Table 1.** Number of celiac disease (CD) cases by time of seroconversion to CD autoimmunity (CDA).

| Seroconversion stage (mo). |  | no. of cases | Mean mo. of seroconversion (IQR) |
| --- | --- | --- | --- |
| Early | 0-18 mo. | 42 | 18.5 (16.1-21.4) |
| Mid | 24-36 mo. | 54 | 29.2 (27.0-33.0) |
| Late | 36-48 mo. | 41 | 42.0 (37.2-43.4) |

**Figure 1.**
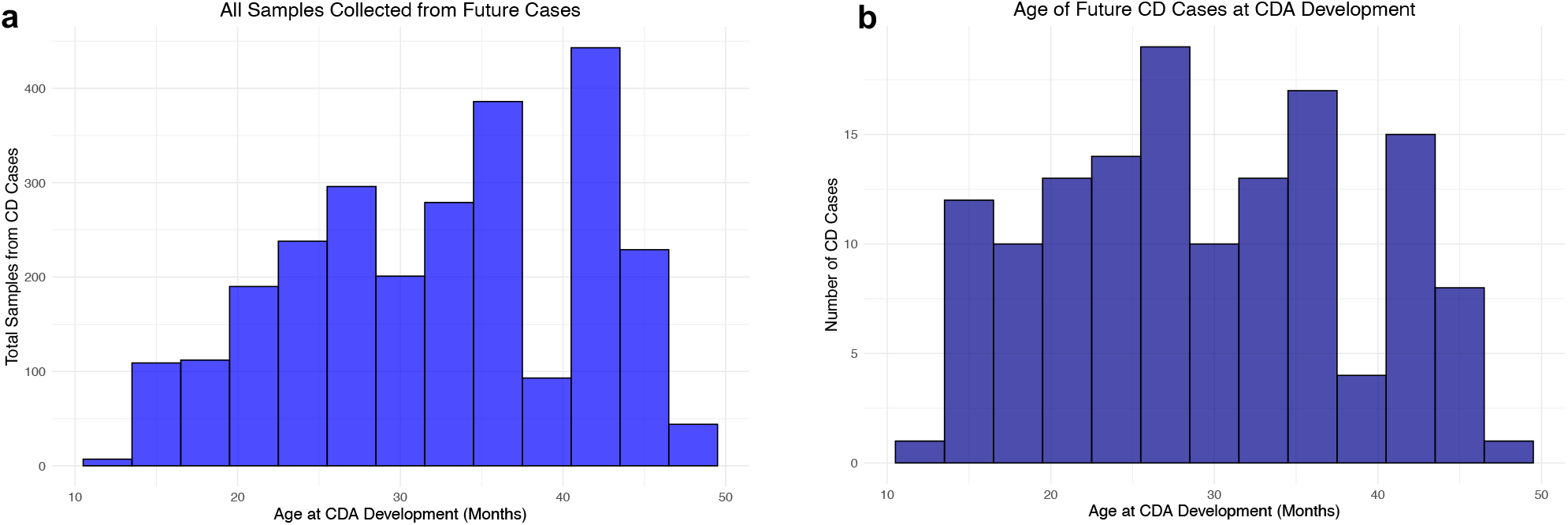
Number of stool samples from CD cases and number of CD cases available for this study as a function of the age in months at time of CDA seroconversion.

DNA was extracted from 8,473 stool samples and sequenced using Illumina targeting the V4 region of the 16S rRNA gene, identifying 1,325 bacterial operational taxonomic units (OTUs), clustered at 97% similarity. To minimize the impact of extreme values, abundances within the 99.9% percentile were retained, removing only the top 0.1%. This approach ensures that rare or low-abundance bacteria are not excluded, maximizing ability to detect subtle but potentially meaningful differences.

### Longitudinal sampling reveals CD microbiome differences preceding seroconversion

To detect microbial abundance differences, the global case-control cohort (all cases combined) was first analyzed, with subsequent stratified analysis based on the child’s age at seroconversion (≤24 months; “early seroconverters”; 24-36 months: “mid seroconverters”; 36-48 months: “late seroconverters”).

Significant microbiome differences were observed in the critical period just before seroconversion regardless of time of birth (Fig. 2a-e). This was observed while accounting for expected microbiome shifts during early childhood, key confounders of CD risk and microbiome composition were balanced using propensity score matching across future cases and controls.

**Figure 2.**
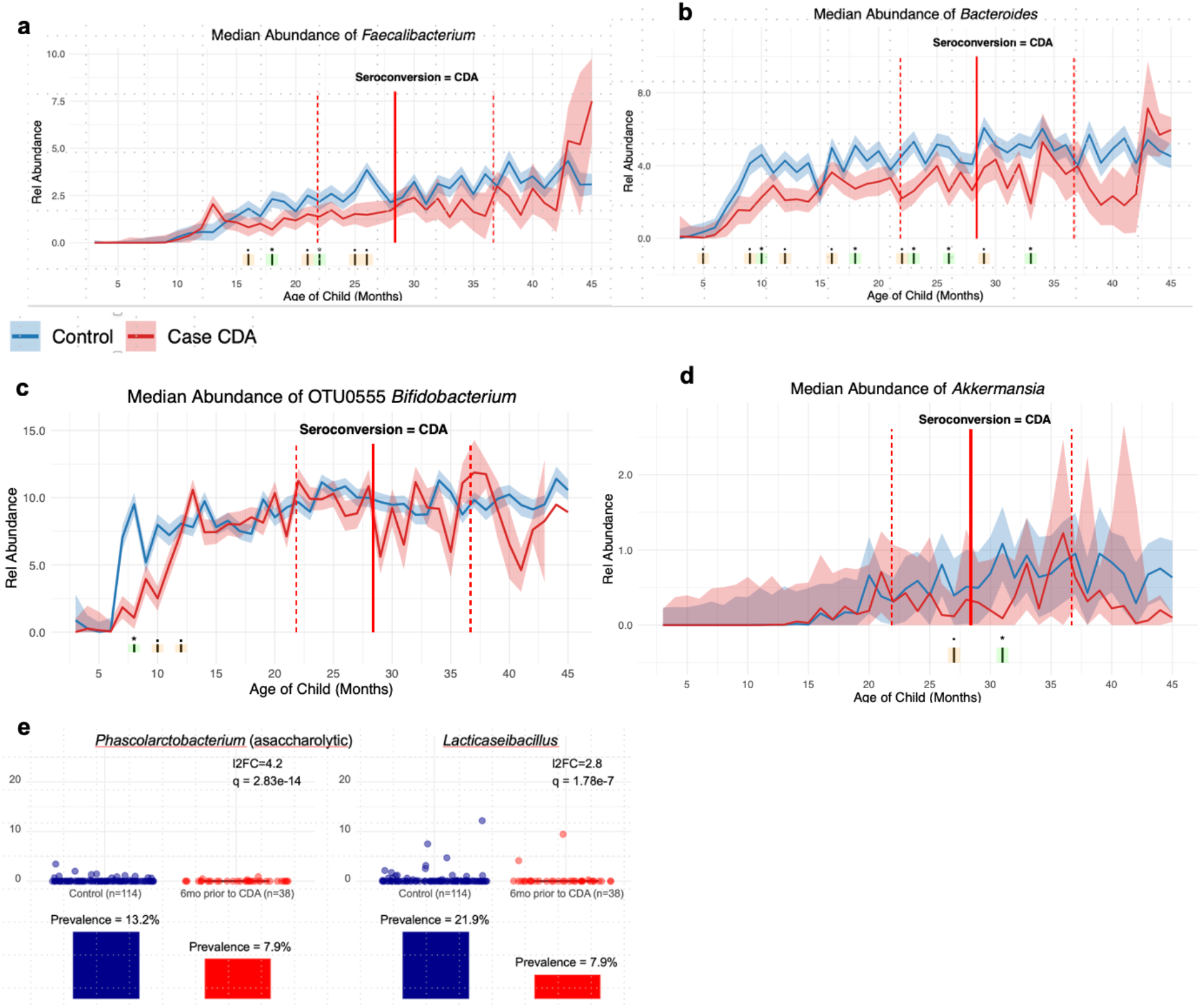
Taxonomic differences up to 48 months of age. **(a-d)** Significant increases in genus-level relative abundance betweens in controls compared to D cases over the first four years of life, separated by timepoint. The relative abundance of specific taxa across time is shown in cases (red) and controls (blue). Vertical lines above the timelines on the x-axis represent specific timepoints where cases and controls differed significantly with an adjusted p-value of <0.01 (yellow) or <0.05 (green). **(e)** Relative abundance of *Lacticaseibacillus* and *Phascolarctobacterium*, six months before seroconversion. Propensity score matching was applied to adjust for a variety of factors (sex, maternal education, parity, age at gluten introduction, breastfeeding duration, and age at stool collection). Stool samples collected between 3 and 18 months of age were pooled. For cases, samples collected six months prior to seroconversion were analyzed, with a random two age-matched controls randomly selected for each case.

After matching, no significant differences remained between groups for any of these factors: sex (p=0.645), maternal education (p=0.933), parity (p=1.0), age at gluten introduction (p=0.659), total breastfeeding duration (p=0.403), or duration of exclusive breastfeeding (p=0.332). All stool samples collected in these individuals between 3 and 18 months of age were pooled. For future cases, samples collected six months prior to the visit at which seroconversion were then moved forward for analysis. For each case, two age-matched controls were randomly selected.

Bacterial genera significantly higher in controls across all periods prior to seroconversion included *Faecalibacterium* (Fig. 2a), *Bacteroides* (Fig. 2b), *Bifidobacterium* (Fig. 2c), and *Akkermansia* (Fig. 2d). *Lacticaseibacillus* and *Phascolarctobacterium* are depleted in cases six months before seroconversion, independent of child age and key confounders. Two of the strongest genus-level differences in abundance (Fig. 2e) were identified in *Lacticaseibacillus* (l2FC = 2.8, q = 1.78e-7) and *Phascolarctobacterium* (l2FC = 4.3, q = 2.83e-14). Only 7.9% of cases (at 6 months prior to CDA) had at least 0.1% *Lacticaseibacillus*, compared to 21.9% of controls. At this same threshold of abundance, *Phascolarctobacterium* was also half as prevalent in CD cases.

#### Cases with early seroconversion of celiac disease autoimmunity (CDA)

In the CDA-phase analysis of all early seroconverters (CDA 13-24 months), several OTU and genus-level differences were noted between case and control children across time. For example, OTU1138 *Collinsella*, likely *Collinesella aerofaciens*, based on sequencing alignment with NCBI BLAST, spiked higher in cases just before and persisted within the window of seroconversion (Fig. 3b). *Erysipelotrichaceae UCG 003*, (d) *Ruminococcus*, (e) *Enterococcus*, and (f) *Turicibacter* across time.

**Figure 3.**
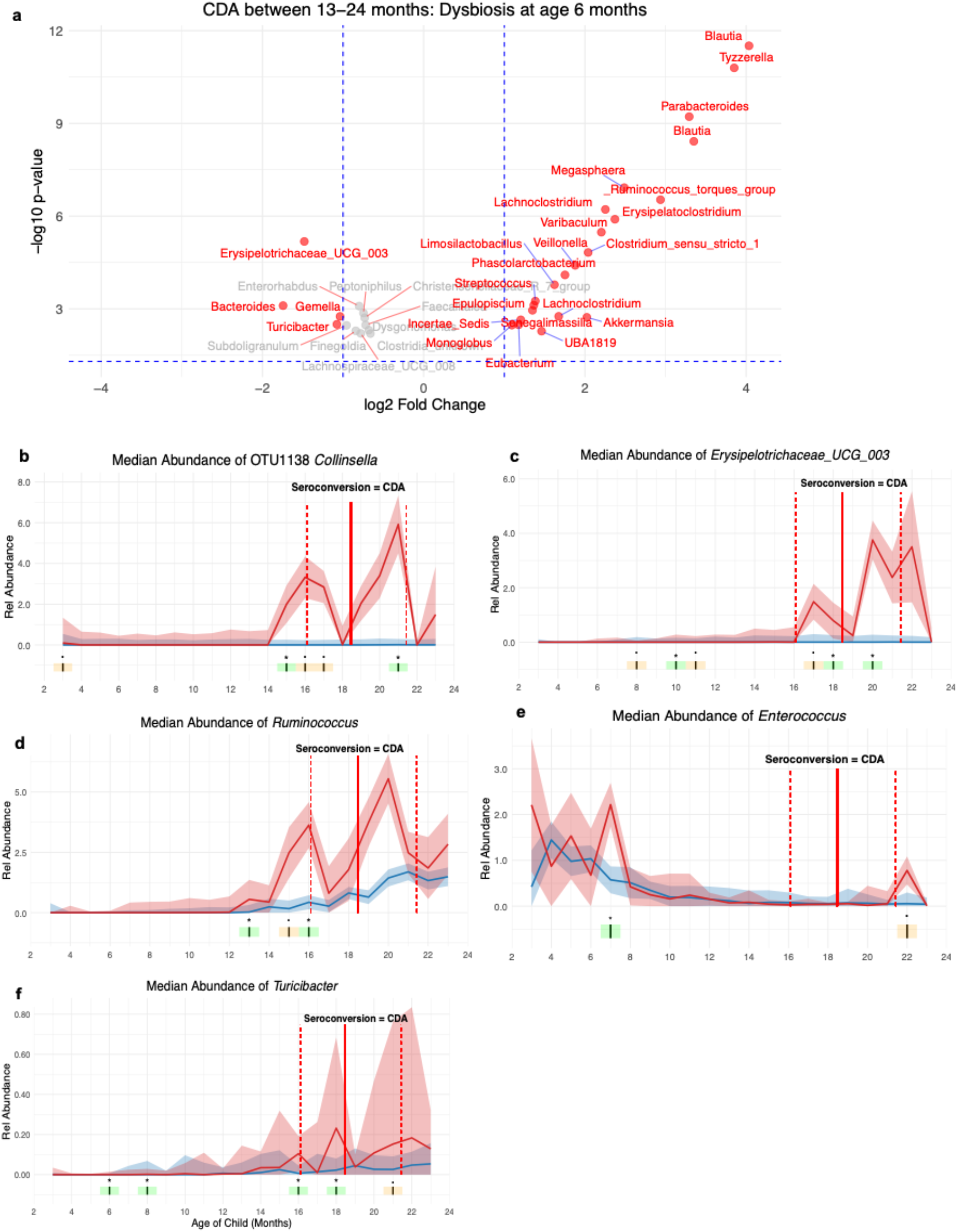
Cases with early seroconversion of celiac disease autoimmunity (CDA) Dysbiosis that occurs at 6 months of age, comparing controls and CD cases seroconverting between 13-24 months of age (early seroconverters), (a) with red dots and labels corresponding to taxa significant after FDR correction. Key differential microbial abundancdifferences between early seroconverters (CDA ≤24 months) and controls, including (b) *Collinsella* OTU1138, (c) *Erysipelotrichaceae UCG 003*, (d) *Ruminococcus*, (e) *Enterococcus*, and (f) *Turicibacter* across time. Vertical lines above timelines on the x-axis represent specific timepoints where cases and controls differed significantly with an adjusted p-value of <0.01 (yellow) or <0.05 (green) (b-f).

#### Cases with mid and late seroconversion of celiac disease autoimmunity (CDA)

During the mid seroconversion time frames, *Faecalibacterium* and certain Clostridia are significantly higher in controls and cases, respectively (Figs. 4). At both mid and late seroconversion time periods, there are several time points where *Bifidobacterium* and *Faecalibacterium* are significantly higher than controls (Figs. 5) while others were higher in cases (Fig. 6). At the late seroconversion period, certain *Bacteroides* are higher at specific time points in controls compared to cases. (Fig. 5)

**Figure 4.**
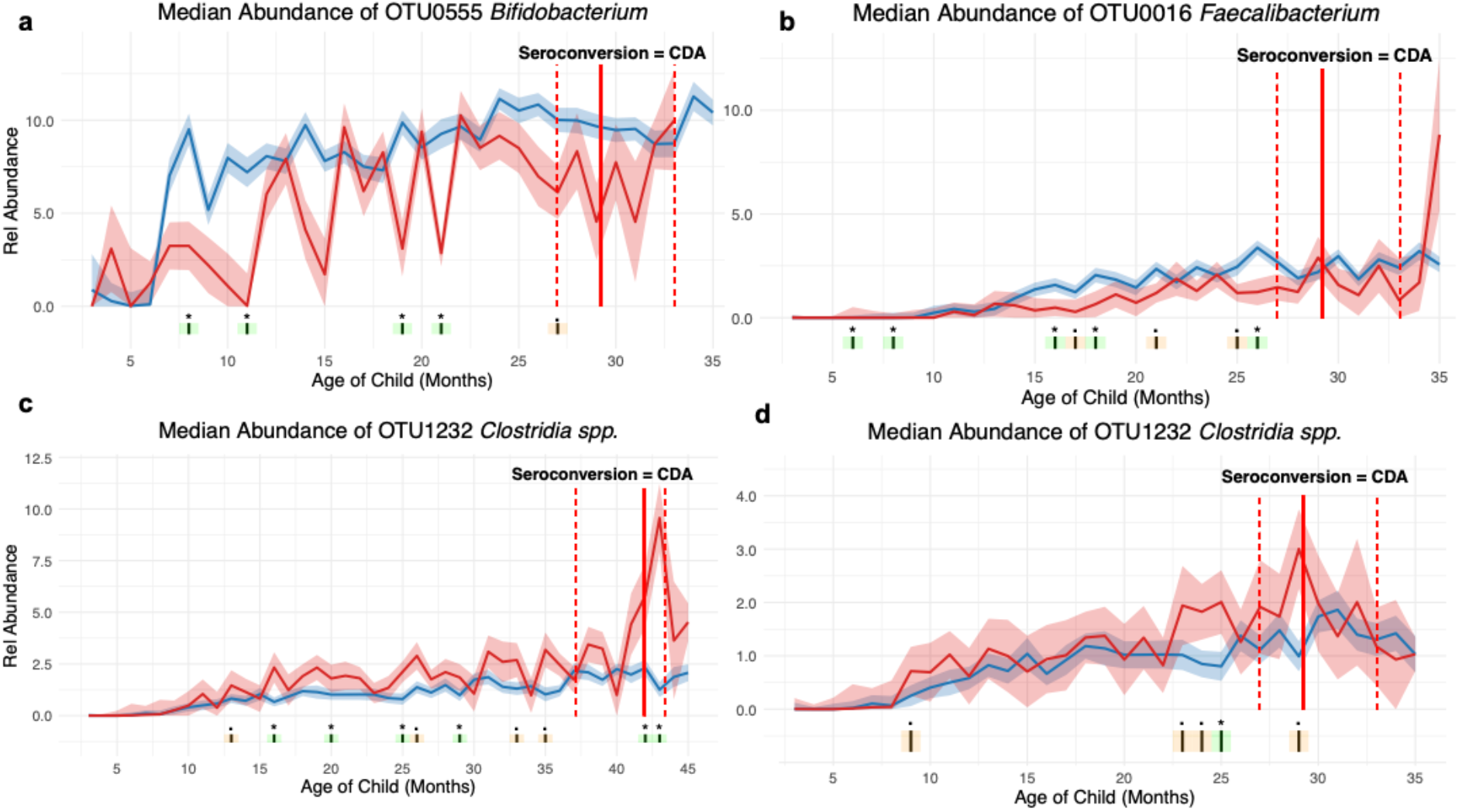
Cases with mid-seroconversion of celiac disease autoimmunity (CDA) Key microbial abundance differences across time between **(a-d)** controls and CD cases who seroconverted between 24-36 months (mid seroconverters) across time. Vertical lines above the timelines on the x-axis represent specific timepoints where cases and controls differed significantly with an adjusted p-value of <0.01 (yellow) or <0.05 (green).

**Figure 5.**
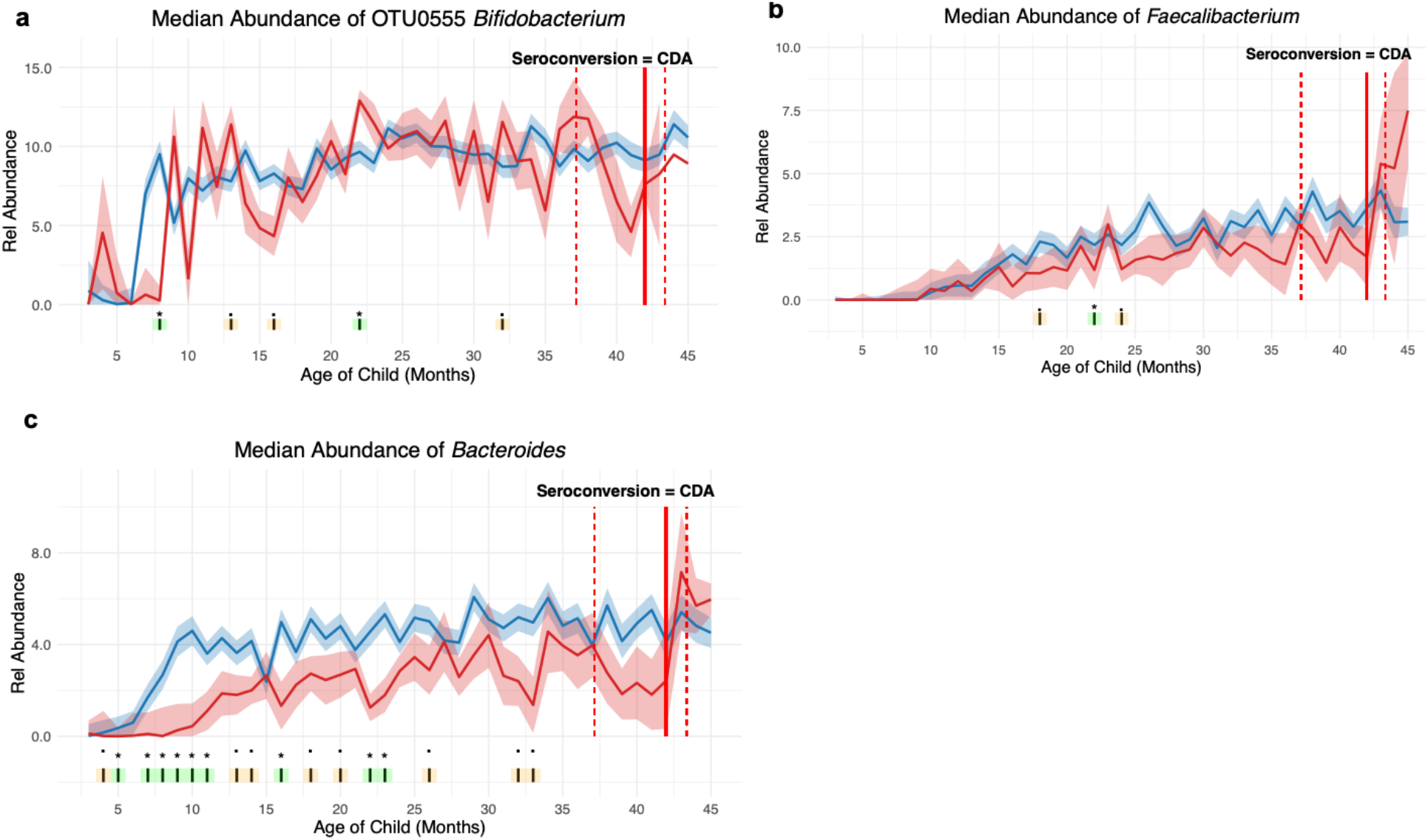
Cases with late seroconversion of celiac disease autoimmunity (CDA) Key microbial abundance differences **(a-c)** across time higher in controls than CD cases who seroconverted between 36-48 months (late seroconverters) across time. Vertical lines above the timelines on the x-axis represent specific timepoints where cases and controls differed significantly with an adjusted p-value of <0.01 (yellow) or <0.05 (green).

**Figure 6.**
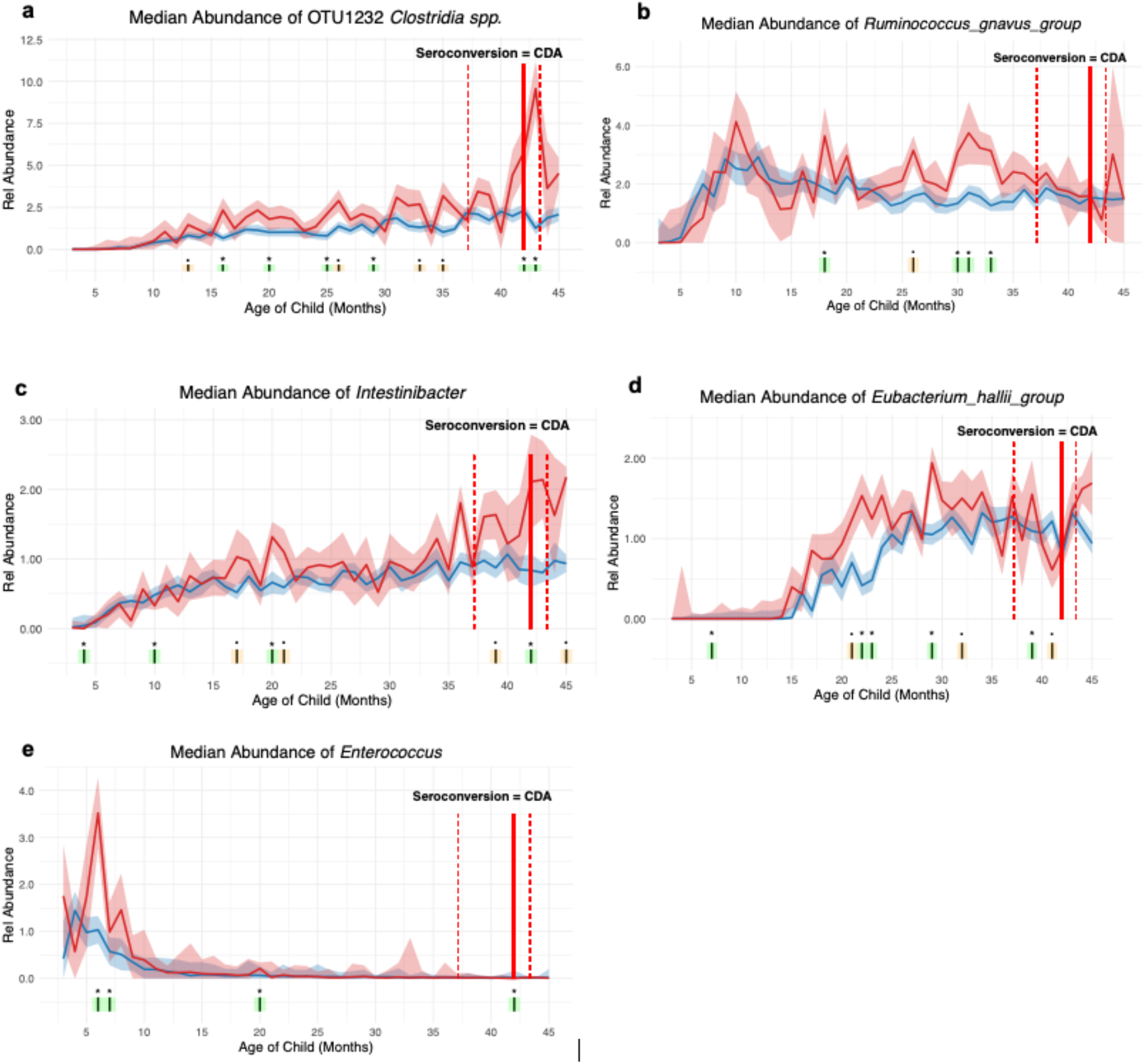
Cases with late seroconversion of celiac disease autoimmunity (CDA) Key microbial abundance differences **(a-d)** across time higher in CD case than in controls in those who seroconverted between 36-48 months (late seroconverters) across time. Vertical lines above the timelines on the x-axis represent specific timepoints where cases and controls differed significantly with an adjusted p-value of <0.01 (yellow) or <0.05 (green).

## DISCUSSION

Across the entire seroconversion period, CD cases were significantly deficient in important bacteria bacterial genera that contribute to gut health including *Bifidobacterium, Akkermansia*, and *Faecalibacterium. Bifidobacterium* produces antioxidants and converts lactate to acetate^20,21^. *Bifidobacterium* has also been shown in cross sectional studies to be in lower relative abundance in subjects with celiac disease^22,23^. As a result, strains in this bacterial genus are being tested as probiotics to reduce celiac disease symptoms^24–26^. (Alternatives to a gluten-free diet are necessary as this diet changes alone do not necessarily resolve all issues^27^. *Akkermansia* is commonly associated with gut health compared to subjects with a variety of gut ailments^28^ including celiac disease^29^. *Faecalibacterium* similarly is considered anti-inflammatory and has been associated with reduced coronary artery disease^30^ and inflammatory bowel disease^31^ as well as playing a modulatory role in immune responses^32^. Here, *Bacteroides* was also higher in controls. Some beneficial strains of Bacteroides have been described as promising probiotics to reduce gut leakiness^33^ and have been shown to increase in children on a gluten-free diet^34^.

The significantly lower prevalence of *Lacticaseibacillus* and *Phascolarctobacterium* in cases six months prior to seroconversion was also striking. Strains of *Lacticaseibacillus* are well characterized for their probiotic, antioxidant, and anti-inflammatory properties^35,36^. *Phascolarctobacterium* strains are asaccharolytic^37^ of this genus was recently discovered to adjust innate immunity and reduce obesity in a mouse model^38^. Given these characteristics, it is not surprising that *Phascolarctobacterium* would be more common in those consuming less gluten and avoiding CD.

The gut dysbiosis in those who seroconvert have an abundance of bacteria more highly associated with cases than controls. This lack of protective bacteria may be contributing to the early seroconversion in the first two years of life. These harmful bacteria include *Collinsella, Enterococcus*, Erysipelotrichaceae, *Ruminococcus*, and *Turicibacter. Collinsella* is considered pro-inflammatory and associated low fiber intake, obesity, and type 2 diabetes^39,40^. Likely *Collinsella aerofaciens*, based on sequencing alignment by NCBI BLAST, this species is known to produce a pH-responsive lipid immunogen that induces the production of proinflammatory markers, TLR2 and TNF-α, both associated with CD^41^.

In the analysis of mid- and late seroconverters, OTU0192 *Bacteroides* was significantly lower across time, from age five months well into the second year of life. *Bacteroides* can stimulate the immune system and enhance phagocytosis of macrophages^42^. Important to its fitness are mucins. The polysaccharide A (PSA) produced by *Bacteroides fragilis* contributes to T-cell responses and has shown protective properties in mouse models of colitis, correcting immune deficiencies and preventing inflammation^43^, necessary for inducing CD4^+^ and CD8^+^ T cells that control inflammation via IL-10^44^.

*Enterococcus* strains are well known for their antimicrobial resistance^45^ which can lead to their higher abundance when a child is treated for a disease caused by bacteria. Unclassified bacteria in the family *Erysipelotrichaceae* possess GalNAc catabolism pathway and degrades N-acetylgalactosamine, an inmportant component of mucin, a protein required for gut integrity^46^. *Ruminococcus* is abundant in inflammatory bowel disease^47^ and Chron’s disease^48^. And like *Erysipelotrichaceae, Ruminococcus* strains can degrade mucin^49^. *Turicibacter* strains can promote ROS-associated apoptosis^50^. Clostridia were found to be higher in cases that were mid-seroconverters. In a large population study, subjects with CD were twice as likely to have a *Clostridium difficile* infection as those without CD^51^.

Our interpretation of these results is that gut dysbiosis associated with future early childhood CDA and CD can occur very early in life. The severity of the dysbiosis appears to be associated with the timing of serconversion (Fig. 7). Those who seroconvert early have an abundance of bacteria that can lead to inflammation and lessened gut integrity. Late seroconversion appears to be more associated with the reduced abundance of beneficial bacteria rather than harmful bacteria. Notably, these beneficial bacteria were consistently more abundant in controls throughout the first four years of life.

**Figure 7.**
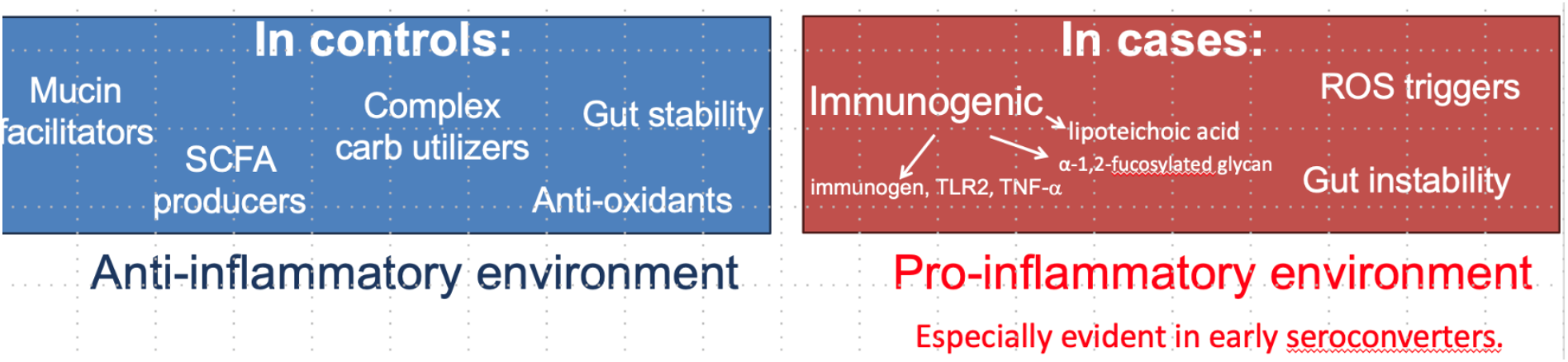
Model for the role of gut bacteria in the development of the inflammatory environment contributing to celiac disease.

## RESOURCE AVAILABILITY

### Materials Availability

This study did not generate new unique reagents.

### Data and Code Availability

Processed microbiome data derived from human samples will be deposited at the time of peer-reviewed publication likely using DRYAD.

### Approvals from Institutional Review Boards

Institutional Review Boards of Colorado’s Colorado Multiple Institutional Review Board, Georgia’s Medical College of Georgia Human Assurance Committee (2004-2010)/Georgia Health Sciences University Human Assurance Committee (2011-2012)/Georgia Regents University Institutional Review Board (2013-2016)/Augusta University Institutional Review Board (2017-present), Florida’s University of Florida Health Center Institutional Review Board, Washington state’s Washington State Institutional Review Board (2004-2012)/Western Institutional Review Board (2013-2019)/WCG IRB (2020-present), and European Ethics Committee Boards of Finland’s Ethics Committee of the Hospital District of Southwest Finland, Germany’s Bayerischen Landesärztekammer (Bavarian Medical Association) Ethics Committee, Sweden’s Regional Ethics Board in Lund, Section 2 (2004-2012)/Lund University Committee for Continuing Ethical Review (2013-present) gave *<u>ethical</u>* approval for this work.

## METHODS

A celiac case control cohort was established as an ancillary study in TEDDY, comprising pure controls (without type 1 diabetes), controls who developed CDA before 48 months but did not develop CD before four years of age, and case CDA who developed CDA before 48 months and eventually CD or had high titer in persistent sample. Controls were followed for a maximum of 48 months in this investigation.

### CD Case-Cohort Design

To combine the flexible advantages of a cohort study with the efficiency of a nested case-control (NCC), a Celiac Case-Cohort has been designed to examine virome and microbiome content of 18,200 stool samples on the subsequent risk of Celiac Disease Autoimmunity (CDA) (i.e. persistent tissue transglutaminase autoantibodies tTGA) leading to a biopsy proven celiac disease (CD) diagnosis (CDA-to-CD). The original Celiac TEDDY cohort consisted of 6555 enrolled children carrying HLA high risk haplo-genotypes homozygous or heterozygous for DR3-DQ2 and DR4-DQ8. Children were screened for CDA and islet autoantibodies (IA) before 4 years of age any children developing IA were excluded (n=423, 6132 remaining). The number of CDA-to-CD cases before 4 years of age was 306/6132 (5%) children with a median (interquartile range) age at CDA onset of 2.3 (1.8 – 3.0) years. An additional 398/6132 children developed CDA and not CD. These children were treated in analysis as a competing CDA group with a low risk for CD in childhood. The case-cohort design differs from an NCC study in that a sub-cohort is selected from the original cohort to use as a representative population. All children in the sub-cohort, including any cases chosen by chance, will be weighted in all regression analysis to represent the original Celiac cohort. (case-cohort Ref to add) A random sample of 561/6132 children (overall sampling fraction = 9.2%) were chosen from the original Celiac TEDDY cohort, and 37/561 CDA-to-CD cases were chosen by chance to serve as time varying controls until before seroconversion. Non CDA-to-CD cases were followed till censoring (dropout or age 48 months, N= 423) or until seroconversion for the competing CDA group that had no subsequent CD diagnosis (n=41).. A statistical power analysis was performed to detect significant hazard ratios (HR) with a 9.1% sub-cohort sampling fraction and a 5% CDA-to-CD case incidence proportion assuming a 1% type-1 error rate (i.e. p-value <0.01 is significant).

### Disease outcome measurements

The definitions of the two outcomes in this work, celiac disease autoimmunity (CDA) and celiac disease, were described previously^52^. Trans-tissue glutaminase IgA and IgG autoantibodies (tTG) were determined in two laboratories and the data harmonized as described previously^52^.

### Stool DNA extraction, 16S rRNA sequencing and analysis

DNA was extracted from 8,473 stools samples using the PowerMag Microbiome DNA isolation kit. Amplication of the V4 region of the 16S rRNA gene and 16S rRNA sequencing using Illumina sequencing (1×150 bp reads)^53^. Classification of reads, statistical analysis, and visualization of the 16S rRNA data was done across time was done using DESeq2^54^ which includes false discovery rate adjustments. Confounders considered in the analysis included sex, material education, parity gluten and breastfeeding.

To account for differences in sequencing depth across samples, using DESeq^54^, we applied a median-of-ratios normalization, which estimates size factors for each feature by dividing each sample’s counts by the geometric mean across all samples. A generalized linear model was fitted for each taxon, modeling the count data using a negative binomial distribution. Estimating dispersions for each feature, biological and technical variability can be accounted for. To improve the reliability of fold change estimates, empirical Bayes shrinkage was employed, especially useful for low-abundance taxa. Wald tests were used to assess statistical significance, and multiple testing correction was applied using the Benjamini-Hochberg procedure to control the false discovery rate (FDR).

## REFERENCES

1. Lebwohl, B., Sanders, D.S., and Green, P.H.R. (2018). Coeliac disease. Lancet 391:70– 81. 10.1016/S0140-6736(17)31796-8.

2. Kuja-Halkola, R., Lebwohl, B., Halfvarson, J., Wijmenga, C., Magnusson, P.K.E., and Ludvigsson, J.F. (2016). Heritability of non-HLA genetics in coeliac disease: a population-based study in 107 000 twins. Gut 65:1793–1798. 10.1136/gutjnl-2016-311713.

3. Lindfors, K., Lin, J., Lee, H.-S., Hyöty, H., Nykter, M., Kurppa, K., Liu, E., Koletzko, S., Rewers, M., Hagopian, W., et al. (2020). Metagenomics of the faecal virome indicate a cumulative effect of enterovirus and gluten amount on the risk of coeliac disease autoimmunity in genetically at risk children: the TEDDY study. Gut 69:1416–1422. 10.1136/gutjnl-2019-319809.

4. TEDDY Study Group (2008). The Environmental Determinants of Diabetes in the Young (TEDDY) Study. Ann N Y Acad Sci 1150:1–13. 10.1196/annals.1447.062.

5. Andrén Aronsson, C., Lee, H.-S., Hård Af Segerstad, E.M., Uusitalo, U., Yang, J., Koletzko, S., Liu, E., Kurppa, K., Bingley, P.J., Toppari, J., et al. (2019). Association of Gluten Intake During the First 5 Years of Life With Incidence of Celiac Disease Autoimmunity and Celiac Disease Among Children at Increased Risk. JAMA 322:514–523. 10.1001/jama.2019.10329.

6. Stahl, M., Koletzko, S., Andrén Aronsson, C., Lindfors, K., Liu, E., Agardh, D., and TEDDY Study Group (2024). Coeliac disease: what can we learn from prospective studies about disease risk? Lancet Child Adolesc Health 8:63–74. 10.1016/S2352-4642(23)00232-8.

7. Hård af Segerstad, E.M., Mramba, L.K., Aronsson, C.A., Uusitalo, U., Virtanen, S.M., Agardh, D., Norris, J.M., Koletzko, S., Rewers, M.J., Toppari, J., et al. (2025). Early Dietary Fiber Intake Reduces Celiac Disease Risk in Genetically Prone Children: Insights from the TEDDY study. Gastroenterology 10.1053/j.gastro.2025.01.241.

8. Cait, A., Cardenas, E., Dimitriu, P.A., Amenyogbe, N., Dai, D., Cait, J., Sbihi, H., Stiemsma, L., Subbarao, P., Mandhane, P.J., et al. (2019). Reduced genetic potential for butyrate fermentation in the gut microbiome of infants who develop allergic sensitization. J Allergy Clin Immunol 144:1638-1647.e3. 10.1016/j.jaci.2019.06.029.

9. Verdu, E.F., Galipeau, H.J., and Jabri, B. (2015). Novel players in coeliac disease pathogenesis: role of the gut microbiota. Nat Rev Gastroenterol Hepatol 12:497–506. 10.1038/nrgastro.2015.90.

10. Sánchez, E., Donat, E., Ribes-Koninckx, C., Fernández-Murga, M.L., and Sanz, Y. (2013). Duodenal-mucosal bacteria associated with celiac disease in children. Appl Environ Microbiol 79:5472–5479. 10.1128/AEM.00869-13.

11. Wacklin, P., Kaukinen, K., Tuovinen, E., Collin, P., Lindfors, K., Partanen, J., Mäki, M., and Mättö, J. (2013). The duodenal microbiota composition of adult celiac disease patients is associated with the clinical manifestation of the disease. Inflamm Bowel Dis 19:934–941. 10.1097/MIB.0b013e31828029a9.

12. Pandey, H., Jain, D., Tang, D.W.T., Wong, S.H., and Lal, D. (2024). Gut microbiota in pathophysiology, diagnosis, and therapeutics of inflammatory bowel disease. Intest Res 22:15–43. doi: 10.5217/ir.2023.00080

13. Buffet-Bataillon, S., Durão, G., Le Huërou-Luron, I., Rue, O., Le Cunff, Y., Cattoir, V. and Bouguen, G. (2025) Gut microbiota dysfunction in Crohn’s disease. Front. Cell. Infect. Microbiol. 15:1540352. doi: 10.3389/fcimb.2025.1540352

14. Kelley, K., Dogru, D., Huang, Q., Yang, Y., Palm, N.W., Altindis, E., and Ludvigsson, J. (2025). Children who develop celiac disease are predicted to exhibit distinct metabolic pathways among their gut microbiota years before diagnosis. Microbiol Spectr 13:e01468–24. 10.1128/spectrum.01468-24

15. Akobeng, A.K., Singh, P., Kumar, M., and Al Khodor, S. (2020). Role of the gut microbiota in the pathogenesis of coeliac disease and potential therapeutic implications. Eur J Nutr 59:3369– 3390. 10.1007/s00394-020-02324-y.

16. Caminero, A., McCarville, J.L., Galipeau, H.J., Deraison, C., Bernier, S.P., Constante, M., Rolland, C., Meisel, M., Murray, J.A., Yu, X.B., et al. (2019). Duodenal bacterial proteolytic activity determines sensitivity to dietary antigen through protease-activated receptor-2. Nat Commun 10:1198. 10.1038/s41467-019-09037-9.

17. Leonard, M.M., Valitutti, F., Karathia, H., Pujolassos, M., Kenyon, V., Fanelli, B., Troisi, J., Subramanian, P., Camhi, S., Colucci, A., et al. (2021). Microbiome signatures of progression toward celiac disease onset in at-risk children in a longitudinal prospective cohort study. Proc Natl Acad Sci (USA) 118:e2020322118. 10.1073/pnas.2020322118.

18. Caminero, A., Galipeau, H.J., McCarville, J.L., Johnston, C.W., Bernier, S.P., Russell, A.K., Jury, J., Herran, A.R., Casqueiro, J., Tye-Din, J.A., et al. (2016). Duodenal Bacteria From Patients With Celiac Disease and Healthy Subjects Distinctly Affect Gluten Breakdown and Immunogenicity. Gastroenterology 151:670–683. 10.1053/j.gastro.2016.06.041.

19. Herrán, A.R., Pérez-Andrés, J., Caminero, A., Nistal, E., Vivas, S., Ruiz de Morales, J.M., and Casqueiro, J. (2017). Gluten-degrading bacteria are present in the human small intestine of healthy volunteers and celiac patients. Res Microbiol 168:673–684. 10.1016/j.resmic.2017.04.008.

20. Averina, O.V., Poluektova, E.U., Marsova, M. V., & Danilenko, V. N. (2021). Biomarkers and Utility of the Antioxidant Potential of Probiotic Lactobacilli and Bifidobacteria as Representatives of the Human Gut Microbiota. Biomedicines 9:1340. 10.3390/biomedicines9101340

21. Talwalkar, A. and Kailasapathy, K. (2003). Metabolic and Biochemical Responses of Probiotic Bacteria to Oxygen. J Dairy Science 86:2537-2546. DOI: 10.3168/jds.S0022-0302(03)73848-X

22. Krishnareddy, S. (2019). The microbiome in celiac disease. Gastroenterol Clin North Am 48:115–126. 10.1016/j.gtc.2018.09.008

23. Matera, M., Guandalini, S. (2024) How the microbiota may affect celiac disease and what we can do. Nutrients 16:1882. 10.3390/nu16121882

24. Norouzbeigi, S., Vahid-Dastjerdi, L., Yekta, R., Sohrabvandi, S., Zendeboodi, F., and Mortazavian, A.M. (2020). Celiac therapy by administration of probiotics in food products: a review. Curr Opin Food Sci 32:58–66. 10.1016/j.cofs.2020.01.005

25. Giorgi, A., Cerrone, R., Capobianco, D., Filardo, S., Mancini, P., Zanni, F., Fanelli, S., Mastromarino, P., and Mosca, L. (2020). A probiotic preparation hydrolyzes gliadin and protects intestinal cells from the toxicity of pro-inflammatory peptides. Nutrients 12:495. 10.3390/nu1202049

26. Mozafarybazargany, M., Khonsari, M., Sokoty, L. et al. (2023). The effects of probiotics on gastrointestinal symptoms and microbiota in patients with celiac disease: a systematic review and meta-analysis on clinical trials. Clin Exp Med 23:2773–2788. 10.1007/s10238-022-00987-x

27. Malamut, G., Cording, S., and Cerf-Bensussan, N. (2019). Recent advances in celiac disease and refractory celiac disease. F1000Res 8:F1000 Faculty Rev–969. 10.12688/f1000research.18701.1

28. Cani, P.D., Depommier, C., Derrien, M. et al. (2022). Akkermansia muciniphila: paradigm for next-generation beneficial microorganisms. Nat Rev Gastroenterol Hepatol 19:625–637. 10.1038/s41575-022-00631-9

29. Roque, A., Zanker, J., Brígido, S. et al. (2024). Dietary patterns drive loss of fiber-foraging species in the celiac disease patients gut microbiota compared to first-degree relatives. Gut Pathog 16:58. 10.1186/s13099-024-00643-7

30. Yang, H.T., Jiang, Z.H., Yang, Y. et al. (2024). Faecalibacterium prausnitzii as a potential Antiatherosclerotic microbe. Cell Commun Signal 22:54. 10.1186/s12964-023-01464-y

31. Cabezas-Cruz A. and Bermúdez-Humarán, L.G. (2024). Exploring the relationship between Faecalibacterium duncaniae and Escherichia coli in inflammatory bowel disease (IBD): Insights and implications, Comp Struct Biotechnol J23:1-9. 10.1016/j.csbj.2023.11.027.

32. Zhang, J., Huang, Y.J., Trapecar, M. et al. (2024). An immune-competent human gut microphysiological system enables inflammation-modulation by Faecalibacterium prausnitzii. npj Biofilms Microbiomes 10:31 10.1038/s41522-024-00501-z

33. Tran, T., Senger, S., Baldassarre, M. et al. (2024). Novel Bacteroides Vulgatus strain protects against gluten-induced break of human celiac gut epithelial homeostasis: a pre-clinical proof-of-concept study. Pediatr Res 95:1254–1264. 10.1038/s41390-023-02960-0

34. Zhang, J., Huang, YJ., Trapecar, M. et al. (2024). An immune-competent human gut microphysiological system enables inflammation-modulation by Faecalibacterium prausnitzii. npj Biofilms Microbiomes 10:31 10.1038/s41522-024-00501-z

35. Bousmaha-Marroki, L., Boutillier, D., Marroki, A. et al. (2021). In Vitro Anti-staphylococcal and Anti-inflammatory Abilities of Lacticaseibacillus rhamnosus from Infant Gut Microbiota as Potential Probiotic Against Infectious Women Mastitis. Probiotics & Antimicro Prot 13:970–981. 10.1007/s12602-021-09755-x

36. Won, G., Choi, SI., Park, N. et al. (2021). In Vitro Antidiabetic, Antioxidant Activity, and Probiotic Activities of Lactiplantibacillus plantarum and Lacticaseibacillus paracasei Strains. Curr Microbiol 78:3181–3191. 10.1007/s00284-021-02588-5

37. Watanabe Y, Nagai F, and Morotomi M. (2012). Characterization of Phascolarctobacterium succinatutens sp. nov., an Asaccharolytic, Succinate-Utilizing Bacterium Isolated from Human Feces. Appl Environ Microbiol 78:511–518. 10.1128/AEM.06035-11

38. Liébana-García, R., López-Almela, I., Olivares, M. et al. (2025). Gut commensal Phascolarctobacterium faecium retunes innate immunity to mitigate obesity and metabolic disease in mice. Nat Microbiol 10.1038/s41564-025-01989-7

39. Gomez-Arango, L.F., Barrett, H.L., Wilkonson, S.A., Callaway, L.K., McIntyre, H.D., Morrison, M., and Nitert Dekker, M. (2018). Low dietary fiber intake increases Collinsella abundance in the gut microbiota of overweight and obses pregmant women. Gut Microbes 9:189–201. 10.1080/19490976.2017.1406584.

40. Candela, M., Biagi, E., Soverini, M., Consolandi, C., Quercia, S., Severgnini, M., Peano, C., Turroni, S., Rampelli, S., Pozzilli, P. et al. (2016). Modulation of gut microbiota dysbioses in type 2 diabetic patients by macrobiotic Ma-Pi 2 diet. Br J Nutr 116:80–93. doi:10.1017/S0007114516001045.

41. Kwon, J., Bae, M., Szamosvári, D., Cassilly, C.D., Bolze, A.S., Jackson, D.R., Xavier, R.J., and Clardy, J. (2023). Collinsella aerofaciens Produces a pH-Responsive Lipid Immunogen. J Am Chem Soc 145:7071–7074. 10.1021/jacs.3c00250.

42. Deng, H., Li, Z., Tan, Y., Guo, Z., Liu, Y., Wang, Y., Yuan, Y., Yang, R., Bi, Y., Bai, Y., et al. (2016). A novel strain of Bacteroides fragilis enhances phagocytosis and polarises M1 macrophages. Sci Rep 6:29401. 10.1038/srep29401.

43. Troy, E.B., and Kasper, D.L. (2010). Beneficial effects of Bacteroides fragilis polysaccharides on the immune system. Front Biosci 15:25–34.

44. Ramakrishna, C., Kujawski, M., Chu, H., Li, L., Mazmanian, S.K., and Cantin, E.M. (2019). Bacteroides fragilis polysaccharide A induces IL-10 secreting B and T cells that prevent viral encephalitis. Nat Commun 10:2153. 10.1038/s41467-019-09884-6.

45. Wei, Y., Palacios Araya, D. and Palmer, K.L. (2024). Enterococcus faecium: evolution, adaptation, pathogenesis and emerging therapeutics. Nat Rev Microbiol 22:705–721. 10.1038/s41579-024-01058-6

46. Zhou, M., Wu, J., Wu, L., Sun, X., Chen, C., and Huang, L. (2024). The utilization of N-acetylgalactosamine and its effect on the metabolism of amino acids in Erysipelotrichaceae strain. BMC Microbiol 24:397. 10.1186/s12866-024-03505-z

47. Nagao-Kitamoto, H. and Kamada, N. (2017). Host-microbial Cross-talk in Inflammatory Bowel Disease”. Immune Network 17:1–12. doi:10.4110/in.2017.17.1.1

48. Henke, M.T. Kenny, D.J., Cassilly, C.D. Vlamakis, H., Xavier, R.J., and Clardy, J. (2019 Ruminococcus gnavus, a member of the human gut microbiome associated with Crohn’s disease, produces an inflammatory polysaccharide, Proc Natl Acad Sci (USA) 116:12672–12677, doi:10.1073/pnas.1904099116

49. Schaus, S.R., Vasconcelos Pereira, G., Luia, A.S., Madlambayan, E., Teerapon, N., Pstrowski, M.P., Jin, C., Henrissat, B., Hansson, G.C., and Martens, E.C.. (2024). Ruminococcus torques is a keystone degrader of intestinal mucin glycoprotein, releasing oligosaccharides used by Bacteroides thetaiotaomicron. mBio 15:e00039–24. 10.1128/mbio.00039-24

50. Lin, T-C, Soorneedi, A., Guan, Y., Tang, Y., Shi,,E., Moore, M.D. and Liu, Z. (2023). Turicibacter fermentation enhances the inhibitory effects of Antrodia camphorata supplementation on tumorigenic serotonin and Wnt pathways and promotes ROS-mediated apoptosis of Caco-2 cells. Front Pharmacol 14:1203087. doi: 10.3389/fphar.2023.1203087

51. Lebwohl, B., Nobel, Y.R., Green, P.H.R., Blaser, M.J., and Ludvigsson J.F. (2017). Risk of Clostridium difficile Infection in Patients With Celiac Disease: A Population-Based Study. Am J Gastroenterol 112:1878–1884. doi: 10.1038/ajg.2017.400.

52. Liu, E., Lee, H, Aronsson, C.A., Hagopian, W.A., Koletzko, S., Rewers, M.J., Eisenbarth, G.S., Bingley, P.J., Bonifacio, E., Simell, V., and Agardh, D. for the TEDDY Study Group. (2014). New England J Med 371:42–49. DOI: 10.1056/NEJMoa1313977

53. Stewart, C.J., Ajami, N.J., O’Brien, J.L. et al. (2018). Temporal development of the gut microbiome in early childhood from the TEDDY study. Nature 562:583–588. 10.1038/s41586-018-0617-x

54. Love, M.I., Huber, W., and Anders, S. (2014). Moderated estimation of fold change and dispersion for RNA-seq data with DESeq2. Genome Biology 15:550. 10.1186/s13059-014-0550-8.

